# Pre-stroke physical activity and Val66Met (rs6265) genotype of *BDNF* gene correlate with the post-stroke cognitive outcome: a prospective cohort study

**DOI:** 10.64898/2026.03.19.26348718

**Authors:** Dariusz Kotlega, Barbara Peda, Agnieszka Zembron-Lacny, Katarzyna Baldy-Chudzik, Edyta Wawrzyniak-Gramacka, Malgorzata Szczuko

**Author notes:** Correspondence author: Dariusz Kotlega.

## Abstract

**Background:** Pre-stroke physical activity may protect against post-stroke cognitive impairment (PSCI) via neuroplastic mechanisms including BDNF signalling. However, evidence in stroke survivors and the modifying role of the *BDNF* Val66Met polymorphism remain limited.

**Methods:** In this prospective single-centre cohort, 97 patients with ischaemic stroke underwent detailed neuropsychological assessment during index hospitalisation and at 6-month follow-up. Pre-stroke activity was quantified as weekly MET-minutes (IPAQ scale). Serum BDNF and Val66Met genotype were measured. Associations between METs and cognitive outcomes (baseline, follow-up) were assessed with Spearman correlations and adjusted multivariable linear regression.

**Results:** Higher pre-stroke METs correlated with superior baseline performance across global cognition (MoCA), verbal learning/recall (CVLT indices), attention and executive measures. After adjustment, CVLT long-delay free recall (LDFR) and TMTA remained significant (β for CVLT LDFR ≈0.000097 per MET-min/week, p < 0.05). At 6 months, METs were associated with multiple CVLT indices (SDCR, SDFR, LDFR) in adjusted models. All associations indicated positive correlation between physical activity and better cognitive outcome. Mean serum BDNF did not correlate with METs (mean 27,261 ± 7,967 pg/mL) and did not differ from controls. Val/Val (GG) carriers of Val66Met genotype of *BDNF* gene performed better cognitively than Met allele carriers, but the genotype did not correlate with serum BDNF level.

**Conclusions:** Greater self-reported pre-stroke physical activity was associated with better early and 6-month cognitive outcomes after ischaemic stroke, whereas peripheral BDNF levels were not correlated. Val66Met polymorphism of *BDNF* gene correlates with the cognitive outcome in stroke survivors but doesn’t enhance BDNF level, indicating effect on activity rather than quantity.

## Introduction

Ischaemic stroke is a leading cause of disability in adults. It is associated with a wide range of post-stroke symptoms that adversely affect quality of life, including speech difficulties, imbalance, falls, sarcopenia, pain, fatigue and cognitive symptoms. Post-stroke cognitive impairment (PSCI) is one of the most common sequelae and occurs in up to 64% of stroke survivors (Saunders et al., 2014). Even low-intensity physical exercise has been shown to exert beneficial effects on global cognition, memory and executive function across different populations (Singh et al., 2025). Available data also indicate that physical activity after stroke is associated with enhanced cognitive performance [Cumming et al., 2012]. Accordingly, it has been recommended that stroke survivors engage in a structured exercise programme at least three times per week for 30–60 minutes per session, amounting to approximately 180 minutes per week, to achieve better cognitive outcomes (Li et al., 2024).

Early cognitive impairment after stroke contributes to reduced levels of physical exercise at six months post-event, which may reflect the fact that cognitive function is required for engagement in most activities, including physical exercise (Viktorisson et al., 2021). Each one-point decrease in Montreal Cognitive Assessment (MoCA) score after stroke has been linked to greater non-adherence to physical activity recommendations [Luzum et al., 2023]. While most studies focus on the relationship between post-stroke physical activity and cognitive outcomes, there is also limited evidence indicating that the level of pre-stroke physical activity correlates with post-stroke cognitive performance (Reinholdsson et al., 2022, Wong et al., 2016, Damsbo et al., 2020, Bovim et al., 2019, Zhu et al., 2025).

The mechanisms underlying the beneficial effects of physical exercise on cognitive outcomes are thought to involve increased cerebral oxygen supply and cerebral perfusion, which stimulate neurogenesis, angiogenesis and synaptic plasticity. In addition, exercise is associated with a reduction in inflammatory processes, mediated by increased expression of neurotrophic factors, reduced oxidative stress and decreased β-amyloid formation (Luzum et al., 2023). It has been suggested that the association between gait speed and subjective memory complaints may be explained by shared risk factors, including cardiovascular disease, diabetes mellitus, dysregulated cortisol profiles, vitamin D deficiency, brain atrophy with reduced hippocampal volume, and increased cerebral β-amyloid deposition (Semba et al., 2020). The pathophysiology of post-stroke cognitive impairment (PSCI) is primarily related to focal structural brain lesions caused by infarction, as well as secondary neuroinflammatory responses that exacerbate neuronal injury. Stroke location is also a critical determinant of PSCI risk (Chen et al., 2023, Schellhorn et al., 2021). Pro-inflammatory processes and ischaemic brain injury are mediated by cytokines and immune cells infiltrating the blood–brain barrier (BBB), together with increased oxidative stress (Zhao et al., 2022, Xia et al., 2023).

Brain-derived neurotrophic factor (BDNF) is a member of the neurotrophin family and plays a key role in axonal regeneration, cognitive modulation, angiogenesis, synaptogenesis and neuroplasticity. It is highly expressed in the hippocampus and exerts its effects primarily through activation of the tropomyosin receptor kinase B (TrkB). BDNF has been identified as a potential mediator of exercise-induced improvements in cognitive function, as both BDNF signalling and physical activity enhance long-term potentiation and thereby support neuroplastic processes (Romero Garavito et al., 2025). Genetic variability within the *BDNF* gene is well established, and several common single nucleotide polymorphisms (SNPs) have demonstrated clinical relevance (Kotlega et al., 2017). Depending on the specific SNP variant, BDNF synthesis in response to physical exercise may differ (Bugge Kambestad et al., 2023), and certain variants have been shown to augment memory performance following exercise interventions (Piepmeier et al., 2020).

One of the most extensively studied polymorphisms, Val66Met (rs6265), has been associated with an increased risk of conversion from mild cognitive impairment (MCI) to Alzheimer’s disease (Xie et al., 2017). However, evidence regarding the role of the Val66Met polymorphism in modulating the relationship between physical exercise and specific cognitive domains, such as attention, information processing speed, executive function, memory and global cognition, remains limited and inconsistent (Liu et al., 2020). Furthermore, the Val66Met polymorphism has been linked to cognitive performance and hippocampal volume, although most studies have been conducted in non-stroke populations, and data in stroke survivors are scarce (Dincheva et al., 2012). To our knowledge, no studies have specifically examined the impact of physical exercise on cognitive performance in stroke survivors in relation to the Val66Met SNP of the *BDNF* gene.

The primary aim of this study was to investigate the association between pre-stroke physical activity (quantified as weekly MET-minutes) and post-stroke cognitive performance at baseline and follow-up in patients with ischemic stroke. Secondary aims were to assess the relationship between pre-stroke physical activity and serum BDNF concentration, and whether the *BDNF* Val66Met polymorphism modifies the associations of physical activity with both serum BDNF and cognitive outcomes. We hypothesised that higher pre-stroke physical activity would be associated with better global and domain-specific cognition after stroke, that greater activity would correlate with higher serum BDNF, and that Met-carrier status would attenuate the positive association between activity and BDNF/cognitive improvement. We also aimed at evaluating the effects of Val66Met genotype of *BDNF* gene on cognitive functions in stroke survivors, irrespectively from the impact on BDNF. This study seeks to clarify the extent to which lifestyle factors and neurobiological markers jointly shape cognitive outcomes following cerebrovascular events.

## Material and methods

### Subjects

In this prospective study, we enrolled 97 patients with a diagnosis of ischaemic stroke. All participants were recruited during their hospital stay in the Neurology Department of a district hospital in Poland. The primary inclusion criterion was an acute cerebral ischaemic stroke, diagnosed on the basis of clinical presentation and confirmed by brain imaging (MRI or CT). Routine laboratory investigations and standard stroke management were provided in line with current guidelines. Stroke was defined as a clinical syndrome of sudden onset with focal or global cerebral dysfunction lasting ≥24 hours or resulting in death, attributable to a vascular cause [Sacco et al., 2013, Adams et al., 2007]. Patients were excluded if neuroimaging demonstrated intracerebral haemorrhage. Additional exclusion criteria were active malignancy, autoimmune disease, clinical signs of infection, or body temperature >37.4 °C. Individuals who were unable to provide informed consent due to impaired consciousness or severe speech disturbance were also excluded. At follow-up, the number of patients available for neuropsychological reassessment ranged from 28 to 43, owing to non-attendance at scheduled visits. The control group comprised 33 stroke-free, age- and sex-matched individuals recruited from local university clubs for older adults.

The protocol of the study was approved by the Ethics Committee in Zielona Gora, Poland (decision number 08/73/2017, Feb 27, 2017). The study was conducted in accordance with the Declaration of Helsinki. All patients signed a written informed consent form to participate in the study.

### Neuropsychological Assessment

Neuropsychological testing was carried out by a trained neuropsychologist experienced in the assessment of cognitive disorders in stroke. The cognitive tests were performed during the index hospitalisation, 7 days after admission, and again at the 6-month follow-up visit. The number of patients reassessed at follow-up was lower than at baseline due to non-attendance.

Participants completed the 21-item Beck Depression Inventory–II (BDI-II), rating each item from 0 to 3 to indicate how they had felt over the previous two weeks. The instrument maps onto DSM-IV diagnostic criteria for major depressive disorder; total scores span 0–63, with larger values denoting greater severity of depressive symptoms (Beck et al., 1996, Reeves et al., 2012).

Cognitive functioning was assessed with a neuropsychological test battery that included the California Verbal Learning Test, Second Edition (CVLT-II), a measure of verbal learning, episodic memory, recognition memory, and attention. We analysed the following CVLT-II indices: List A, List B, short-delay free recall (SDFR), short-delay cued recall (SDCR), long-delay free recall (LDFR), long-delay cued recall (LDCR), and semantic clustering (Delis et al., 2000, Yi et al., 2011). Raw CVLT-II scores were converted into standard ten (sten) scores for all indices. The sten is a normalized psychological scale (mean = 5.5, SD = 2) divided into ten intervals; this transformation, routinely used in clinical neuropsychology, permits comparison across individuals and aids interpretation in correlational analyses. Higher sten values indicate superior cognitive performance.

Short-term verbal memory, attention, and verbal working memory were evaluated using the Forward and Backward Digit Span Tests (FDST and BDST), subtests drawn from the Wechsler Adult Intelligence Scale and the Wechsler Memory Scale (Woods et al., 2011, Fink et al., 2014).

The Trail Making Test (TMT), consisting of Parts A and B, was used to evaluate psychomotor processing speed, sustained attention, visuospatial scanning, working memory, cognitive flexibility, and set-shifting. Test performance was quantified as the time required to complete each part, recorded in seconds, with longer completion times reflecting poorer cognitive performance (Talarowska et al., 2012, Cahn et al., 1995).

Inhibitory control and the ability to adjust behaviour in response to changing task demands were examined using the Stroop Colour and Word Test (SCWT), also known as the Stroop Dots Trial A and B (SDT A and SDT B). An experimental Polish adaptation derived from the original version of the test was administered (Scarpina et al., 2017).

Verbal fluency was evaluated using a Verbal Fluency Test administered under two conditions: semantic (category) fluency and phonemic (letter) fluency. Semantic fluency was assessed with the Verbal Fluency Test–Animals (VFT-A) and the Verbal Fluency Test–Fruits and Vegetables (VFT-F&V), whereas phonemic fluency was measured using the Verbal Fluency Test–Letter “K” (VFT-K) and the Verbal Fluency Test–Letter “P” (VFT-P). A Polish adaptation of the original instrument was employed. This test battery was used to examine executive functions, expressive language abilities, memory processes, and processing speed (Cahn et al., 1995).

Neuropsychological outcomes labelled with the suffix “follow up” denote measurements collected at the 6-month follow-up assessment. With the exception of the Trail Making Test (TMT) and Stroop Dots Test (SDT), higher scores across all instruments indicated superior performance within the respective cognitive domains.

The Montreal Cognitive Assessment (MoCA) was administered as a brief and clinically practical screening instrument to assess global cognitive functioning, encompassing attention, memory, orientation, language, visuospatial skills, and abstract reasoning (Zietemann et al., 2018).

Pre-stroke physical activity was assessed with the use of short form of International Physical Activity Questionnaire (IPAQ) scale. This self-reported questionnaire consists of brief items assessing the frequency and duration of walking, moderate, and vigorous physical activity, and sitting time over the past seven days, providing an overall physical activity estimate suitable for surveillance. Results are calculated into total energy expenditure per week (MET min/week) (Balboa-Castillo et al., 2023).

### BDNF and Val66Met polymorphism analysis

The plasma BDNF concentration was evaluated by using the R&D Systems ELISA kit (USA). The intra-assay coe□cient of variation (CV) was <5%, and the detection limit was estimated at 20 pg/mL for the used kit.

Genomic DNA was isolated from blood using the GeneJET Genomic DNA Purification Kit (Thermo Scientific) according to the manufacturer’s protocol for mammalian cells. DNA concentration and purity were assessed spectrophotometrically using a NanoPhotometer (Implen). Purified DNA was stored at −80 °C until further analysis. BDNF Val66Met genotyping was performed using the ARMS (tetra primer amplified refractory mutation system) method (ARMS-PCR). The method utilizes the specificity of four primers in a single PCR reaction, which allows for the amplification of two allele-specific products (253 and 201 bp) and the entire region (401 bp) as an internal control, as previously described (Sheikh et al., 2010). The first set of primers, P1 and P2 [P1 (forward): 5’-CCT ACA GTT CCA CCA GGT GAG AAG AGT G-3’; P2 (reverse): 5’-TCA TGG ACA TGT TTG CAG CAT CTA GGT A-3’] amplifies the 401-bp rs6265 region of the BDNF gene. The pair in the second set served to identify G/A substitution, where primer P3 is specific for the G allele [P3: 5’-CTG GTC CTC ATC CAA CAG CTC TTC TAT AAC-3’] and primer P4 is specific for the A allele [P4: 5’-ATC ATT GGC TGA CAC TTT CGA ACC CA-3’]. The PCR amplification mixture in a volume of 25 µL contained: buffer solution for (Thermo Scientific, Waltham, MA, USA), 2.5 mM MgSO4 (Promega, Madison, WI, USA), 0.5 mM of each dNTP (Promega), 0.2 µM of each of four primers (Genomed, Poland), 1 U of Platinum Taq DNA Polymerase (Thermo Scientific), and 3 µL of DNA template. The PCR products were separated by electrophoresis on a 3% agarose gel and stained with ethidium bromide.

### Statistical methods

Statistical analyses were performed using standard methods for continuous and categorical data. Continuous variables (e.g., age, serum BDNF concentrations, MET minutes per week) were inspected for distributional properties using the Shapiro-Wilk test and visual plots. Depending on normality and homogeneity of variances, between-group comparisons of continuous variables (main cohort vs. control group) were conducted using either Welch’s t-test (for approximately normally distributed variables with unequal variances) or the nonparametric Mann-Whitney U test (for skewed distributions). Categorical variables (sex, *BDNF* Val66Met genotypes) were summarized as counts and percentages and compared between cohorts using chi-square tests of independence; for small expected counts, Fisher’s exact test was additionally reported. *BDNF* Val66Met genotype in the patient cohort was coded numerically. In the control cohort, genotype strings were normalized and mapped to the same three-level categorical variable. For some analyses, genotypes were further collapsed into Val/Val (GG) versus Met-carriers (GA+AA) to form a binary variable. Serum BDNF levels were compared between the main MET cohort and the control group overall, and genotype distributions were compared between cohorts using 3×2 (GG, GA, AA × cohort) and 2×2 (GG vs Met-carrier × cohort) contingency tables. Age and sex were compared between cohorts to characterize sample differences: age as a continuous variable (Welch’s t-test) and sex as a binary categorical variable (chi-square test). Correlation analyses between MET (minutes per week) and serum BDNF were conducted using Spearman’s rank correlation coefficient in the overall sample and stratified by *BDNF* Val66Met genotype (GG, GA, AA) as well as by genotype grouping (GG vs Met-carriers). Two-sided p-values were used throughout, and a significance level of α = 0.05 was adopted. Analyses were performed using Python (version 3.11.6).

## Results

The demographic characteristic is presented in table 1.

**Table 1.** Demographic characteristic of study group.

| Variable | Value | n |
| --- | --- | --- |
| Age (years) | 63.5 ± 12.1 | 97 |
| Sex (Male), n (%) | 43 (44.3) | 97 |
| BMI | 29 ± 5.1 | 97 |
| CRP (mg/L) | 3.08 ± 4.3 | 97 |
| LDL (mg/dL) | 111.63 ± 45.61 | 97 |
| HDL (mg/dL) | 52.55 ± 15.62 | 97 |
| Hypertension, n (%) | 84 (85.6%) | 97 |
| Diabetes/IFG, n (%) | 47 (48.4%) | 97 |
| Current Smoking, n (%) | 40 (41.2%) | 97 |
| Atrial Fibrillation, n (%) | 10 (10.3%) | 97 |
| Previous Stroke/TIA, n (%) | 11 (11.3%) | 97 |

**Table 2.** Association between baseline cognitive functions and MET value (min/week) in study group.

| Outcome | n | Spearman's rho | p- value |
| --- | --- | --- | --- |
| CVLT LDFR (sten) | 63 | 0.449 | < 0.001 |
| MOCA | 97 | 0.335 | < 0.001 |
| TMTA time | 97 | -0.332 | < 0.001 |
| BDST | 97 | 0.311 | < 0.005 |
| TMTA ten | 97 | -0.293 | < 0.005 |
| Stroop2 time | 97 | -0.268 | < 0.05 |
| VFT-Letter K | 97 | 0.265 | < 0.05 |
| VFT-Fruits and Vegetables | 97 | 0.2593 | < 0.05 |
| TMTB time | 97 | -0.247 | < 0.05 |
| FDST | 97 | 0.215 | < 0.05 |
| CVLT SDFR (sten) | 63 | 0.263 | < 0.05 |
| CVLT List A | 96 | 0.205 | < 0.05 |
| VFT-Letter P | 97 | 0.195 | NS |
| CVLT List A (sten) | 63 | 0.209 | NS |
| CVLT LDCR (sten) | 62 | 0.204 | NS |
| CVLT SDCR (sten) | 63 | 0.196 | NS |
| Stroop1 time | 97 | -0.155 | NS |
| VFT-Animals | 97 | 0.145 | NS |
| TMTB ten | 97 | -0.142 | NS |
| Beck | 97 | -0.108 | NS |
| CVLT List B | 96 | 0.0796 | NS |
| CVLT List B (sten) | 63 | -0.0713 | NS |
| CVLT semantic clustering (sten) | 96 | 0.0106 | NS |

### Baseline cognitive assessments

Analysis of baseline cognitive performance in relation to pre-stroke physical activity, quantified as weekly MET values, demonstrated several significant associations. Higher levels of physical activity were positively correlated with performance on the CVLT List A, CVLT SDFR (sten), Forward Digit Span Test (FDST), Verbal Fluency Test (VFT) for fruits and vegetables, VFT letter K, Backward Digit Span Test (BDST), Montreal Cognitive Assessment (MoCA), and CVLT LDFR (sten).

Conversely, higher physical activity levels were negatively correlated with completion times on the Trail Making Test Part B (TMT-B), Stroop Test Part 2, Trail Making Test Part A (sten), and Trail Making Test Part A completion time, indicating better performance with increasing physical activity. No statistically significant associations were observed for the remaining cognitive measures.

After multivariate adjustment for age, sex and body mass index, CVLT LDFR (sten) remained significantly associated with weekly MET values (p < 0.05; β = 0.000097; 95% CI: 0.000027–0.000167; R^2^ = 0.12). This finding indicates that each additional 1,000 MET-min/week was associated with an approximate 0.097-point increase in CVLT LDFR (sten) score.

Trail Making Test Part A (sten) also remained significantly associated with physical activity, showing an inverse relationship (partial p < 0.05; β = −0.00034; 95% CI: −0.00069 to 0.000005; R^2^ = 0.18). The corresponding regression plots are presented in Figures 1 and 2.

**Figure 1.**
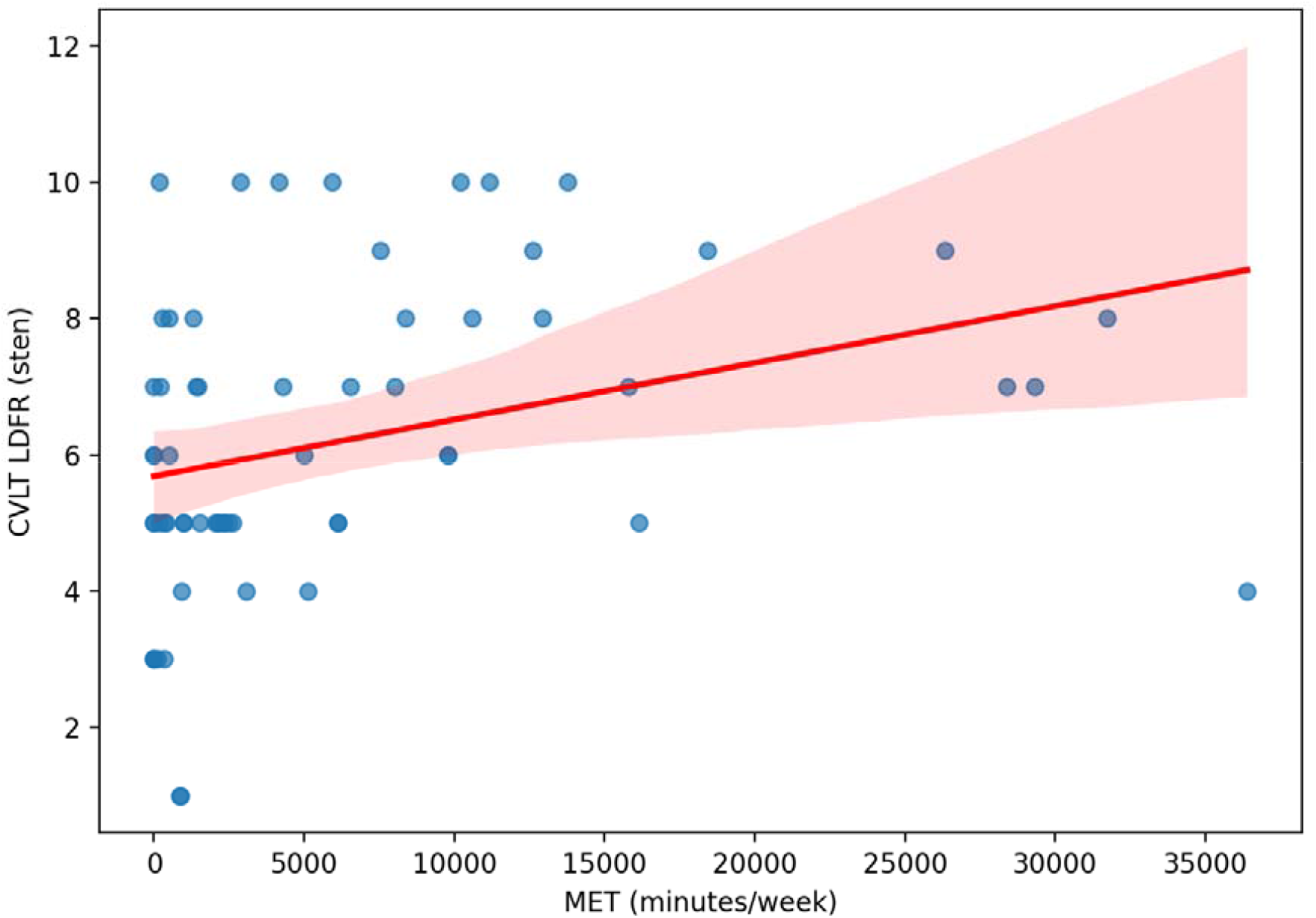
The scatter plot of association between baseline CVLT LDFR (sten) and MET value.

**Figure 2.**
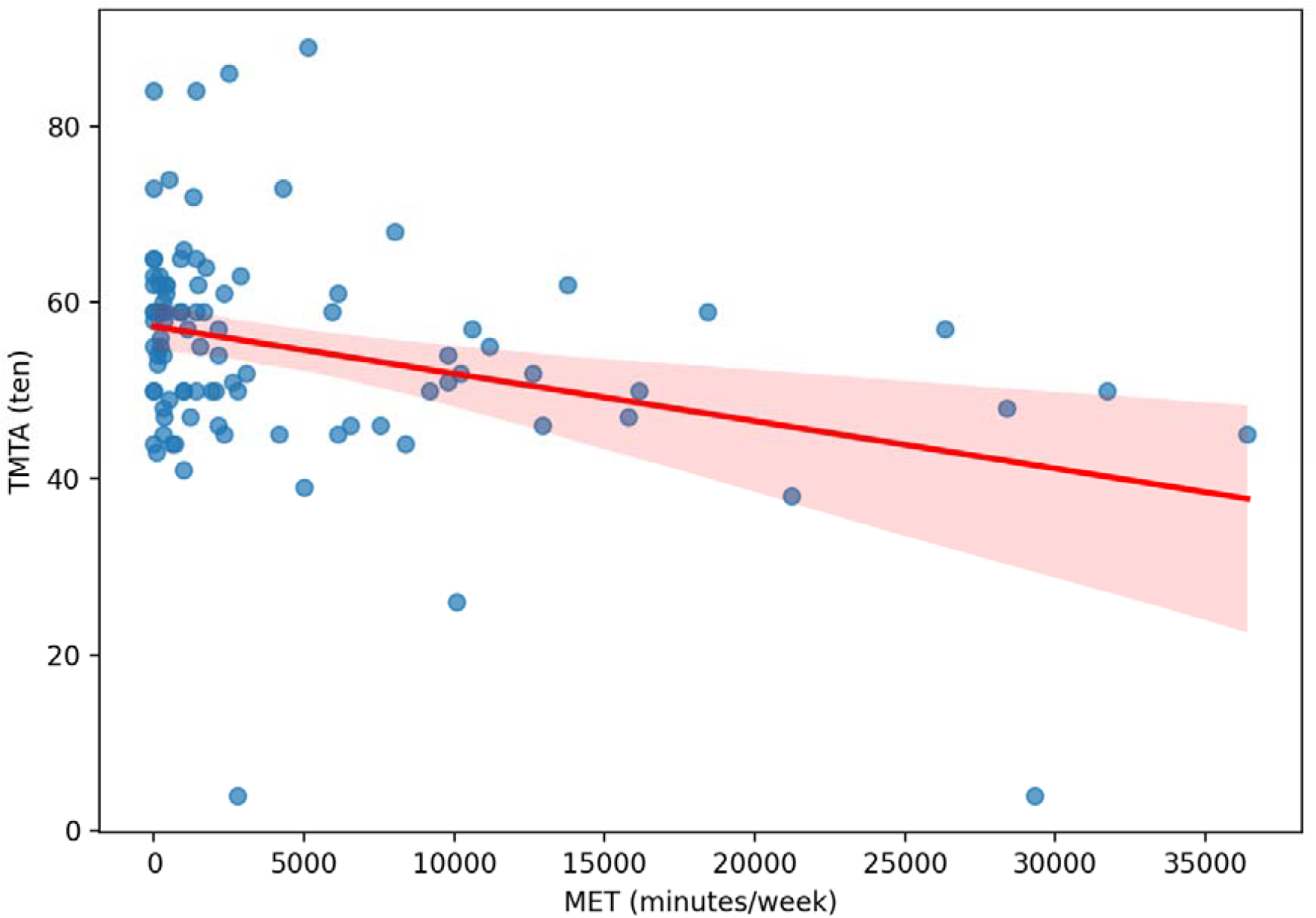
The scatter plot of association between baseline TMTA (ten) and MET value.

### Follow up cognitive assessments

Table 3 summarises the associations between follow-up cognitive test performance and weekly MET values. Higher levels of physical activity were positively correlated with performance on the Verbal Fluency Test (VFT) letter K, CVLT LDFR (sten), VFT fruits and vegetables, and CVLT SDCR (sten). In contrast, higher MET values were negatively correlated with completion times on the Trail Making Test Parts A and B, indicating better performance with greater physical activity. No statistically significant associations were observed for the remaining follow-up cognitive measures.

**Table 3.** Association between follow up cognitive functions and MET value (min/week) in study group.

| **Outcome** | **n** | **Spearman's rho** | **p-value** |
| --- | --- | --- | --- |
| TMTB time follow up | 40 | -0.375 | < 0.05 |
| TMTA time follow up | 40 | -0.363 | < 0.05 |
| CVLT SDCR (sten) follow up | 26 | 0.434 | < 0.05 |
| VFT-Fruits and Vegetables follow up | 33 | 0.374 | < 0.05 |
| CVLT LDFR (sten) follow up | 26 | 0.396 | < 0.05 |
| VFT-Letter K follow up | 34 | 0.343 | < 0.05 |
| VFT-Animals follow up | 33 | 0.329 | NS |
| CVLT SDFR (sten) follow up | 26 | 0.369 | NS |
| VFT-Letter P follow up | 34 | 0.286 | NS |
| Beck follow up | 40 | -0.261 | NS |
| CVLT LDCR (sten) follow up | 26 | 0.313 | NS |
| CVLT List A follow up | 26 | 0.299 | NS |
| CVLT List A (sten) follow up | 26 | 0.297 | NS |
| TMTA (ten) follow up | 40 | -0.208 | NS |
| Stroop1 time follow up | 32 | -0.199 | NS |
| BDST follow up | 38 | 0.172 | NS |
| FDST follow up | 39 | 0.153 | NS |
| CVLT semantic clustering (sten) follow up | 26 | 0.152 | NS |
| Stroop2 time follow up | 31 | -0.115 | NS |
| CVLT List B (sten) follow up | 26 | -0.123 | NS |
| CVLT List B follow up | 26 | -0.110 | NS |
| TMTB (ten) follow up | 40 | -0.067 | NS |

Multivariable analysis confirmed significant associations between weekly MET values and follow-up performance on the CVLT subtests, including CVLT SDCR (sten), CVLT SDFR (sten) and CVLT LDFR (sten). Specifically, for CVLT SDCR (sten), the association remained significant (p < 0.05; β = 0.00011; 95% CI: 0.0000101–0.000211; R^2^ = 0.249). Similarly, CVLT SDFR (sten) demonstrated a significant positive association with physical activity (p < 0.05; β = 0.0000991; 95% CI: 0.00000821–0.000190; R^2^ = 0.242). Finally, CVLT LDFR (sten) also remained significantly associated with MET values (p < 0.05; β = 0.00011; 95% CI: 0.0000991–0.000219; R^2^ = 0.271).

### Associations between BDNF, Val66Met of BDNF gene, and MET values

The mean serum BDNF concentration in the study group was 27,261.43 ± 7,966.91 pg/mL (n = 97). No statistically significant correlation was observed between weekly MET values and serum BDNF concentrations (p > 0.05).

The genotype distribution of the *BDNF* Val66Met polymorphism was as follows: GG in 69 participants (71.1%), GA in 23 participants (23.7%), and AA in 5 participants (5.1%). Serum BDNF concentrations did not differ significantly across genotype groups (Mann–Whitney U test, p > 0.05). Spearman rank correlation analyses between MET values and serum BDNF concentrations stratified by genotype revealed no significant associations in the GG (ρ = 0.078, p > 0.05) or GA groups (ρ = −0.375, p > 0.05). A strong negative correlation was observed in the AA genotype group (ρ = −0.90, p < 0.05); however, this finding should be interpreted with caution due to the very small number of participants with this genotype (n = 5).

Serum BDNF concentrations were comparable between the study group (27,261.43 ± 7,966.91 pg/mL; n = 97) and the control group (26,350 ± 4,968 pg/mL; n = 33), with no statistically significant difference detected (Mann–Whitney U test, p = 0.33). Similarly, the distribution of *BDNF* Val66Met genotypes did not differ between the study and control groups (χ^2^ test, p = 0.397).

A significant difference in age distribution was observed between the study and control groups (63.56 ± 12.13 years vs. 71.18 ± 4.69 years, respectively; Welch’s t-test, p < 0.01). In contrast, sex distribution did not differ between groups, with 43 males in the study group and 14 males in the control group (χ^2^ test, p = 1.0).

The analysis of the impact of the *BDNF* Val66Met polymorphism on cognitive performance is presented in Table 4. Significant associations were observed for the Montreal Cognitive Assessment (MoCA), CVLT semantic clustering (sten), and CVLT List A. Comparisons between GG (Val/Val) genotype carriers and combined Met-allele carriers (GA + AA) were performed using the Mann–Whitney U test. Individuals with the GG (Val/Val) genotype demonstrated significantly better performance on these measures than Met-allele carriers. No statistically significant differences were observed for the remaining cognitive tests (p > 0.05).

**Table 4.** The comparison between Val66Met genotypes of BDNF gene in regards to cognitive functions.

| Variable | Genotype | n | Mean ± SD | p-value |
| --- | --- | --- | --- | --- |
| MOCA | GG | 69 | $24.75 \pm 3.36$ | < 0.05 |
| MOCA | AA+GA | 28 | $23.21 \pm 3.54$ | |
| CVLT semantic clustering (sten) | GG | 68 | $5.62 \pm 2.52$ | < 0.005 |
| CVLT semantic clustering (sten) | AA+GA | 28 | $4.28 \pm 1.90$ | |
| CVLT List A | GG | 68 | $46.79 \pm 10.01$ | < 0.05 |
| CVLT List A | AA+GA | 28 | $42.5 \pm 7.94$ | |

In the adjusted linear regression model, only CVLT semantic clustering (sten) remained statistically significant, with AA/GA carriers scoring on average 1.32 sten units lower than GG homozygotes (β = −1.32; 95% CI: −2.29 to −0.34; p = 0.008).

In the analysis of the impact of the *BDNF* Val66Met polymorphism on follow-up cognitive performance, no statistically significant associations were observed. Comparisons between GG homozygotes and combined Met-allele carriers (AA + GA) using the Mann–Whitney U test did not reveal any significant differences (p > 0.05).

## Discussion

In this prospective cohort study of stroke patients, we examined the associations between pre-stroke physical activity, quantified as weekly MET values, and post-stroke cognitive performance. Baseline cognitive assessments revealed positive correlations between physical activity and CVLT List A, CVLT SDFR (sten), Forward Digit Span Test (FDST), Verbal Fluency Test (VFT) for fruits and vegetables, VFT letter K, Backward Digit Span Test (BDST), Montreal Cognitive Assessment (MoCA), and CVLT LDFR (sten). Conversely, negative correlations were observed with Trail Making Test Part B (TMT-B) time, Stroop Test Part 2 time, TMTA (sten) and TMTA completion time. No statistically significant associations were detected for the remaining cognitive measures. These findings indicate that higher levels of self-reported pre-stroke physical activity are associated with better cognitive performance in the early post-stroke phase, spanning multiple domains including global cognition, memory (verbal learning and recall), attention, working memory, processing speed, and executive function. After multivariate adjustment, the associations with CVLT LDFR (sten) and TMTA (sten) remained statistically significant, highlighting their independent relationship with pre-stroke physical activity.

We also examined the associations between pre-stroke physical activity and cognitive performance at six months post-stroke. Positive correlations were observed for VFT letter K, CVLT LDFR (sten), VFT fruits and vegetables, and CVLT SDCR (sten), whereas negative correlations were found with TMTA and TMTB completion times, indicating better performance with higher physical activity levels. No significant associations were detected for the remaining follow-up cognitive measures. Multivariable analysis confirmed significant associations for CVLT SDCR (sten), CVLT SDFR (sten), and CVLT LDFR (sten), reinforcing the relationship between higher pre-stroke physical activity and superior cognitive outcomes. Overall, these findings are consistent with the baseline assessments, supporting a robust trend linking greater physical activity to improved post-stroke cognitive performance. In a previous study evaluating the impact of physical activity in the 48 hours preceding stroke, a positive correlation with cognitive outcome was also observed. However, cognitive performance in that study was assessed solely using the MoCA test (Reinholdsson et al., 2022). Another study assessed the impact of the total amount of physical activity during the year preceding stroke and found that higher activity levels were associated with a reduced risk of developing post-stroke dementia within 3 to 6 months after the event (Wong et al., 2016). An analysis of physical activity during the seven days preceding stroke demonstrated that more active participants exhibited better cognitive performance. Cognitive function was assessed using the Symbol Digit Modalities Test (SDMT) and the Mini-Mental State Examination (MMSE) at one- and six-months post-stroke (Damsbo et al., 2020). Available evidence demonstrates considerable heterogeneity in methodology across studies. In the present study, we employed the validated International Physical Activity Questionnaire (IPAQ) alongside an extensive battery of neuropsychological assessments conducted by an experienced, specialised clinical psychologist. To date, this represents the most comprehensive evaluation of post-stroke cognitive performance in relation to physical activity.

Although no significant correlation was observed between weekly MET values and serum BDNF concentrations, it should be emphasized that post-stroke cognitive impairment is attenuated by physical activity through multiple molecular mechanisms. Exercise can enhance cerebral blood flow (CBF), improve cerebral hemodynamics, and increase oxygen delivery to the brain. It also promotes neuroplasticity, including synaptic plasticity, neurogenesis, and recruitment of alternative neural pathways to compensate for damaged regions. Brain-derived neurotrophic factor, released in response to increased physical activity, plays a central role in these processes by binding to its high-affinity receptor, tropomyosin receptor kinase B (TrkB), and activating downstream signalling cascades such as MAPK/ERK and PI3K/Akt. Physical activity also modulates neuroinflammatory pathways, including the NLRP3 inflammasome, which mediates cytokine activation. Exercise-induced suppression of NLRP3 activation can reduce M1 microglial activity and enhance BDNF-mediated neuroprotection. Beyond inflammation, physical activity supports neuronal energy metabolism and reactive oxygen species (ROS) management. Peroxisome proliferator-activated receptor gamma coactivator 1-alpha (PGC-1α), a key regulator of mitochondrial biogenesis, is upregulated, while antioxidant enzymes such as superoxide dismutase (SOD) and glutathione peroxidase (GPx) are increased. Activation of the Nrf2/ARE pathway further reduces oxidative stress, protecting neurons from cellular damage (Shui et al., 2024, Zhu et al., 2025, Souza et al., 2022, De Assis et al., 2024).

In addition to cognitive assessments, we measured baseline serum BDNF levels and genotyped participants for the Val66Met single nucleotide polymorphism of the *BDNF* gene. No significant association was observed between pre-stroke MET values and BDNF concentrations, and BDNF levels did not differ according to genotype. Rank correlation analyses suggested that physical activity might influence BDNF in a genotype-dependent manner. However, the only statistically significant correlation was observed in participants with the AA genotype of Val66Met, a group comprising only five individuals, precluding definitive conclusions. We also found no differences in BDNF levels or genotype distribution between the study and control groups. While physical exercise is generally associated with increased BDNF, prior studies in post-stroke cohorts have reported variable results, including no effect of exercise on BDNF, which aligns with our findings despite being somewhat unexpected (Maguire et al., 2023). Other studies indicate that high-intensity physical activity can elevate BDNF levels in stroke survivors, whereas moderate-intensity exercise may not produce the same effect (Boyne et al., 2019, Hsu et al., 2021).

In our study, *BDNF* Val/Val (GG) carriers demonstrated superior cognitive performance compared with Met-allele carriers (AA + GA), particularly in global cognition (MoCA), verbal learning (CVLT List A), and semantic organization during learning (CVLT semantic clustering). Notably, no *BDNF* genotype was associated with higher serum BDNF levels, suggesting that the Val66Met polymorphism may influence BDNF activity rather than its circulating concentration, as proposed by previous studies (Brown et al., 2020, de Assis et al., 2022). The precise role of the Val66Met genotype in modulating the neurotrophic effects of BDNF warrants further investigation. Evidence from other populations supports these observations. In healthy individuals, Met allele carriers exhibited reduced stability of information stored in iconic memory compared with Val/Val homozygotes, indicating that Val/Val individuals retain information in iconic memory for longer periods (Beste et al., 2011). Among patients with bipolar disorder, Met carriers performed significantly worse than Val/Val individuals on the Wisconsin Card Sorting Test (WCST) (Rybakowski et al., 2006). Similarly, in a community sample of elderly White individuals, Val/Val homozygotes outperformed both Val/Met heterozygotes and Met/Met homozygotes on delayed recall and an alphabet-coding task (ACT), a measure of processing speed (Miyajima et al., 2008). Another study reported that Met carriers performed worse than controls on delayed recall tasks, with performance declining further as recall intervals increased (Goldberg et al., 2008). In post-stroke cohorts, lower cognitive performance has been consistently associated with the presence of the Met allele of the *BDNF* Val66Met polymorphism, aligning with our findings (Han et al., 202, Kim et al., 2012). A key distinction of our study is that it represents, to date, the most extensive analysis of post-stroke cognitive outcomes in relation to *BDNF* genotype.

This study has several limitations that constrain the interpretation and generalisability of the findings. The observational, single-centre design precludes causal inference and is susceptible to residual confounding despite multivariable adjustment. Important variables such as detailed premorbid cognitive status and stroke lesion location were not uniformly controlled for. Pre-stroke physical activity was assessed by retrospective self-report on short-form of IPAQ scale, which is vulnerable to recall and reporting bias and lacks the objectivity of device-based measures (e.g., accelerometer). The sample size reduces statistical power for an analysis, especially the follow-up neuropsychological reassessments, which included only 28-43 subjects. The AA genotype subgroup comprised only five individuals, which was underpowered. Peripheral serum BDNF was measured at a single time point, while circulating BDNF may not reliably reflect central nervous system levels or dynamic exercise-related changes, and single measurements increase susceptibility to biological and assay variability. Finally, potential medication effects, rehabilitation exposures and lifestyle changes after stroke were not fully captured and may have influenced follow-up cognitive trajectories.

## Conclusions

In conclusion, this study provides evidence that higher levels of pre-stroke physical activity are associated with more favourable cognitive performance directly after the acute phase of ischaemic stroke and at 6-month follow-up, particularly in domains of verbal learning, memory consolidation and attentional processing. These associations persisted after adjustment for key demographic variables, supporting the concept of physical activity as a form of cognitive reserve that may mitigate post-stroke cognitive impairment. In contrast, peripheral serum BDNF levels were not related to pre-stroke activity or cognitive outcomes, underscoring the complexity of neurotrophic signalling and the limitations of single time-point peripheral measurements as proxies for central neuroplastic processes. The observed differences in selected memory measures between *BDNF* Val66Met genotypes suggest a potential moderating role of genetic factors in post-stroke cognitive trajectories. The results highlight the clinical relevance of promoting regular physical activity as a modifiable, low-cost strategy that may enhance brain resilience before cerebrovascular events. At the same time, they emphasise the need for larger, longitudinal, multicentre studies incorporating objective physical activity assessment, serial biomarker profiling and detailed neuroimaging to clarify underlying mechanisms and to determine whether targeted exercise interventions can meaningfully reduce the burden of cognitive impairment after stroke.

## Data Availability

All data produced in the present study are available upon reasonable request to the authors

